# Activity of convalescent and vaccine serum against a B.1.1.529 variant SARS-CoV-2 isolate

**DOI:** 10.1101/2021.12.20.21268134

**Authors:** Juan Manuel Carreño, Hala Alshammary, Johnstone Tcheou, Gagandeep Singh, Ariel Raskin, Hisaaki Kawabata, Levy Sominsky, Jordan Clark, Daniel C. Adelsberg, Dominika Bielak, Ana Silvia Gonzalez-Reiche, PSP/PARIS Study Group, Komal Srivastava, Emilia Mia Sordillo, Goran Bajic, Harm van Bakel, Viviana Simon, Florian Krammer

## Abstract

The B.1.1.529 (Omicron) variant of severe acute respiratory syndrome coronavirus 2 (SARS-CoV-2) was identified in November of 2021 in South Africa and Botswana as well as in a sample of a traveler from South Africa in Hong Kong.^1,2^ Since then, B.1.1.529 has been detected in many countries globally. This variant seems to be more infectious than B.1.617.2 (Delta), has already caused super spreader events^3^ and has outcompeted Delta within weeks in several countries and metropolitan areas. B.1.1.529 hosts an unprecedented number of mutations in its spike gene and early reports have provided evidence for extensive immune escape and reduced vaccine effectiveness.^2,4-6^ Here, we investigated the neutralizing and binding activity of sera from convalescent, mRNA double vaccinated, mRNA boosted as well as convalescent double vaccinated and convalescent boosted individuals against wild type, B.1.351 and B.1.1.529 SARS-CoV-2 isolates. Neutralizing activity of sera from convalescent and double vaccinated participants was undetectable to very low against B.1.1.529 while neutralizing activity of sera from individuals who had been exposed to spike three or four times was maintained, albeit at strongly reduced levels. Binding to the B.1.1.529 receptor binding domain (RBD) and N-terminal domain (NTD) was reduced in convalescent not vaccinated but was mostly retained in vaccinated individuals.

---

Severe acute respiratory syndrome coronavirus 2 (SARS-CoV-2) emerged in late 2019 in Wuhan, China and has since then caused the coronavirus disease 2019 (COVID-19) pandemic. While SARS-CoV-2 was antigenically relatively stable during its first few months of circulation, the first antigenically distinct variants, B.1.1.7 (Alpha), B.1.351 (Beta) and P.1 (Gamma), emerged in late 2020. Other variants of interest (VOI) and variants of concern (VOCs) followed. So far, B.1.351 has shown the most antigenic drift in terms of reduction of *in vitro* neutralization rivaled only by B.1.621 (Mu).^7^ B.1.617.2 (Delta), which emerged in early 2020 has been the most consequential variant given the fact that it is more infectious than the viruses circulating in the beginning of the pandemic and also partially escapes neutralization *in vitro*.^8^ B.1.1529 (Omicron) was first detected in South Africa, Botswana and in a traveler from South Africa in Hong Kong.^1,2^ The variant hosts a large number of mutations in its spike protein including at least 15 amino acid changes in the receptor binding domain (RBD). These mutations are predicted to affect most neutralizing antibody epitopes. In addition, B.1.1.529 seems to be fit and highly transmissible^3^ and it rapidly spread across the globe. In fact, within four weeks this variant has outcompeted B.1.617.2 and is now the dominant circulating variant in several countries and urban areas.

Immunity to SARS-CoV-2 in human populations is highly variable and likely differs in individuals with infection induced immunity, double vaccinated individuals, boosted individuals and individuals with hybrid immunity due to the combination of infection followed by vaccination. Understanding residual neutralizing and binding activity against highly antigenically distinct viral variants such as B.1.1.529 in these distinct groups is essential to gauge the level of protection that a specific community has against infection, mild or severe COVID-19.

To address these questions, we determined the loss of *in vitro* neutralizing and binding activity for B.1.1529 in sera from individuals with different levels of immunity (infection, vaccine, hybrid). We included samples from convalescent individuals (N=15), individuals vaccinated twice with BNT162b2 (Pfizer/BioNTech mRNA vaccine, N=10), individuals vaccinated twice with mRNA-1273 (Moderna mRNA vaccine, N=10), individuals vaccinated three times with BNT162b2 (boosted, N=10), individuals vaccinated three times with mRNA-1273 (boosted, N=10), convalescent individuals who got 2 doses of BNT162b2 (N=10), convalescent individuals who got 2 doses of mRNA-1273 (N=10) and finally convalescent individuals who got 3 doses of BNT162b2 (boosted, N=10) (**Figure 1A** and **Supplementary Tables 1 and 2**). First, we tested the *in vitro* neutralizing activity of the sera against wild type SARS-CoV-2 (as a reference for ancestral strains), B.1.351 (as a reference for the most pronounced *in vitro* escape phenotype) and B.1.1.529 (isolated from one of the first cases identified in New York City in late November 2021, **Supplementary Table 3**). The neutralization assay used mimics physiological conditions, since it is performed with authentic SARS-CoV-2 in a multicycle replication setting in which serum/antibody is present at all times akin to the situation in a seropositive individual. Across all 85 samples, the reduction in neutralization for B.1.1.529 was greater than 14.5-fold (the actual fold reduction could not be calculated since many samples were below the limit of detection) (**Figure 1B**). In comparison, there was “only” a four-fold reduction against B.1.351 in the same sample set. In fact, 16.5% of samples lost all neutralizing activity against B.1.1529. When looking at the different groups, we noted that convalescent individuals had lower titers against wild type and B.1.351 with the majority (73.3%) of samples having no measurable neutralizing activity for B.1.1529 (**Figure 1C**). For samples from individuals double vaccinated with BNT162b2 and mRNA-1273, we observed a more than 23-fold and a 42-fold reduction in neutralizing activity respectively (**Figure 1D and E**). However, most individuals retained low but detectable neutralizing activity. Boosted individuals experienced lower reduction with a 7.5-fold drop in neutralization for BNT162b2 boosted individuals and a 16.7-fold reduction in mRNA-1273 boosted individuals (**Figure 1F and G**). Of note, the lower fold change and the higher starting neutralization titers led to considerable residual neutralizing activity in those groups. Convalescent individuals who received two BNT162b2, two mRNA-1272 or three BNT162b2 vaccine doses showed 14-fold, 11-fold and 13-fold drops in B.1.1.529 neutralization (**Figure 1H, I and J**). However all individuals in these groups maintained relatively robust neutralization activity. These data indicate that convalescent individuals greatly benefit from vaccination, an observation that is of significant public health importance.

**Figure 1:**
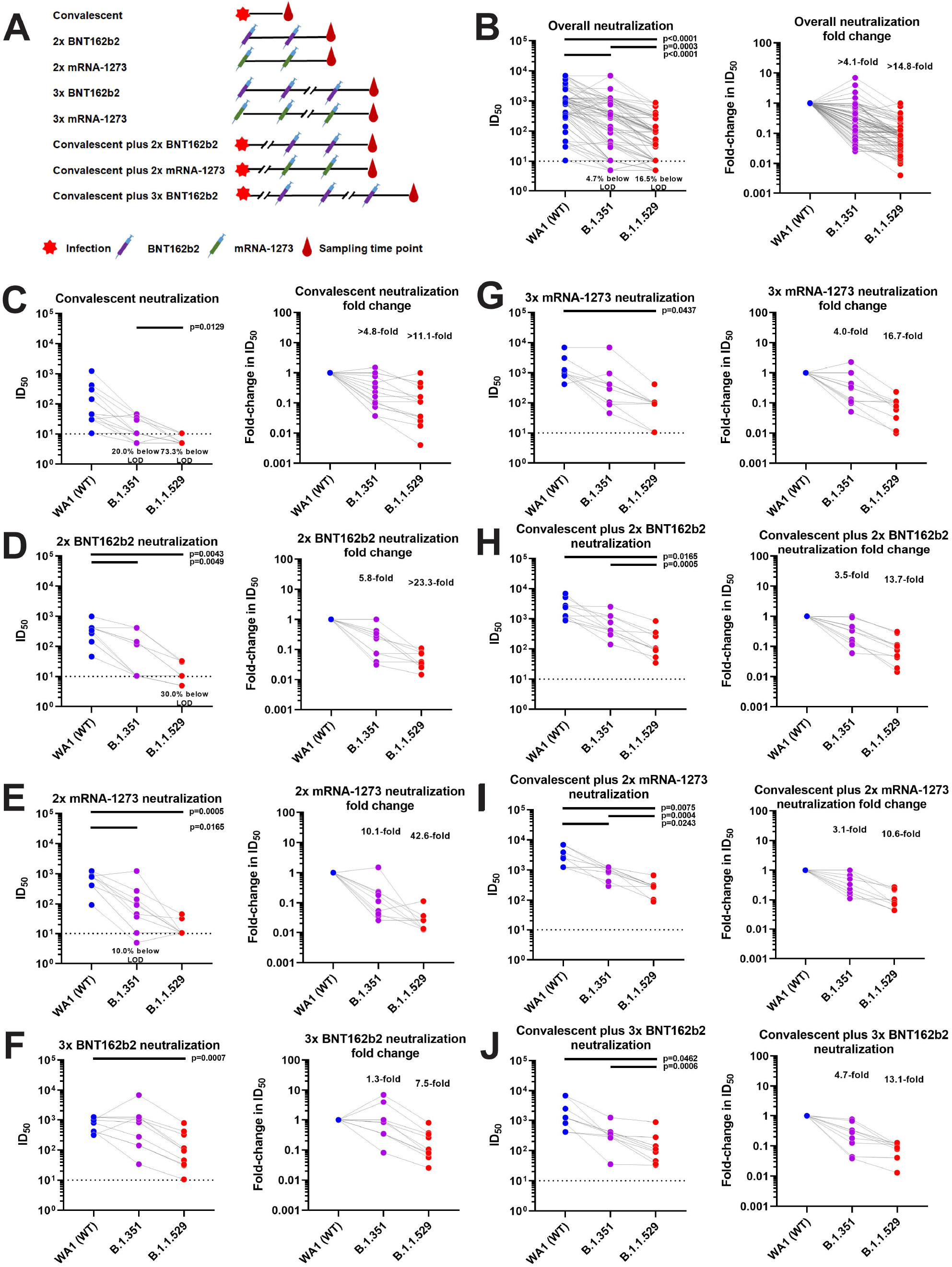
Sera of convalescent and vaccinated individuals have strongly reduced neutralizing activity against B.1.1.529 as compared to wild type SARS-CoV-2. **A** Overview of different exposure groups from whom samples were obtained. Further details are provided in **Supplemental Table 1 and 2. B** shows absolute titers (left) and fold reduction (right) for the combined samples, **C** to **J** shows the different groups. A one-way ANOVA with Tukey’s multiple comparisons test was used to compare the neutralization titers and significant p values (<0.05) are indicated in the figure.

While *in vitro* neutralization is an important antibody function, antibody binding – even in the absence of detectible neutralizing activity – can provide protection through Fc-mediated effector functions. This type of protection has been described in detail for influenza virus^9-11^ but binding antibody titers also represent a correlate of protection for SARS-CoV-2.^12,13^ Furthermore, retained binding to a highly mutated RBD or NTD, even if reduced, indicates that cognate B-cells are present. These B-cells could likely be rapidly recalled during variant infection or variant specific vaccination producing a strong plasmablast response leading to rapid control of viral spread. In addition, B-cells with low affinity binding to antigenically drifted variant proteins may enter lymph notes and engage in germinal center reactions leading to antibodies that may regain neutralizing activity through affinity maturation. To investigate the reduction in binding, we expressed a recombinant RBD of B.1.1.529 and compared binding of this RBD with binding to wild type and B.1.351 RBD (**Figure 2A**). Overall, reduction in binding to B.1.1.529 RBD was much less pronounced than reduction in neutralization (**Figure 2B**). However, this reduction was significantly higher than what we observed here and in the past for B.1.351.^8^ Reduction in binding was most pronounced for convalescent individuals (**Figure 2C**) with a drop of more than 7.5-fold and undetectable reactivity by enzyme linked immunosorbent assay (ELISA) in two thirds of the convalescent individuals who were infected early in the pandemic prior to the circulation of viral variants of concern. In all other groups binding was relatively well maintained with a reduction in binding ranging from a 2.9-fold drop in individuals who had received two vaccinations with mRNA-1273 to a 1.5-fold drop in individuals boosted with BNT162b2 (**Figures 2D to 2J**).

**Figure 2:**
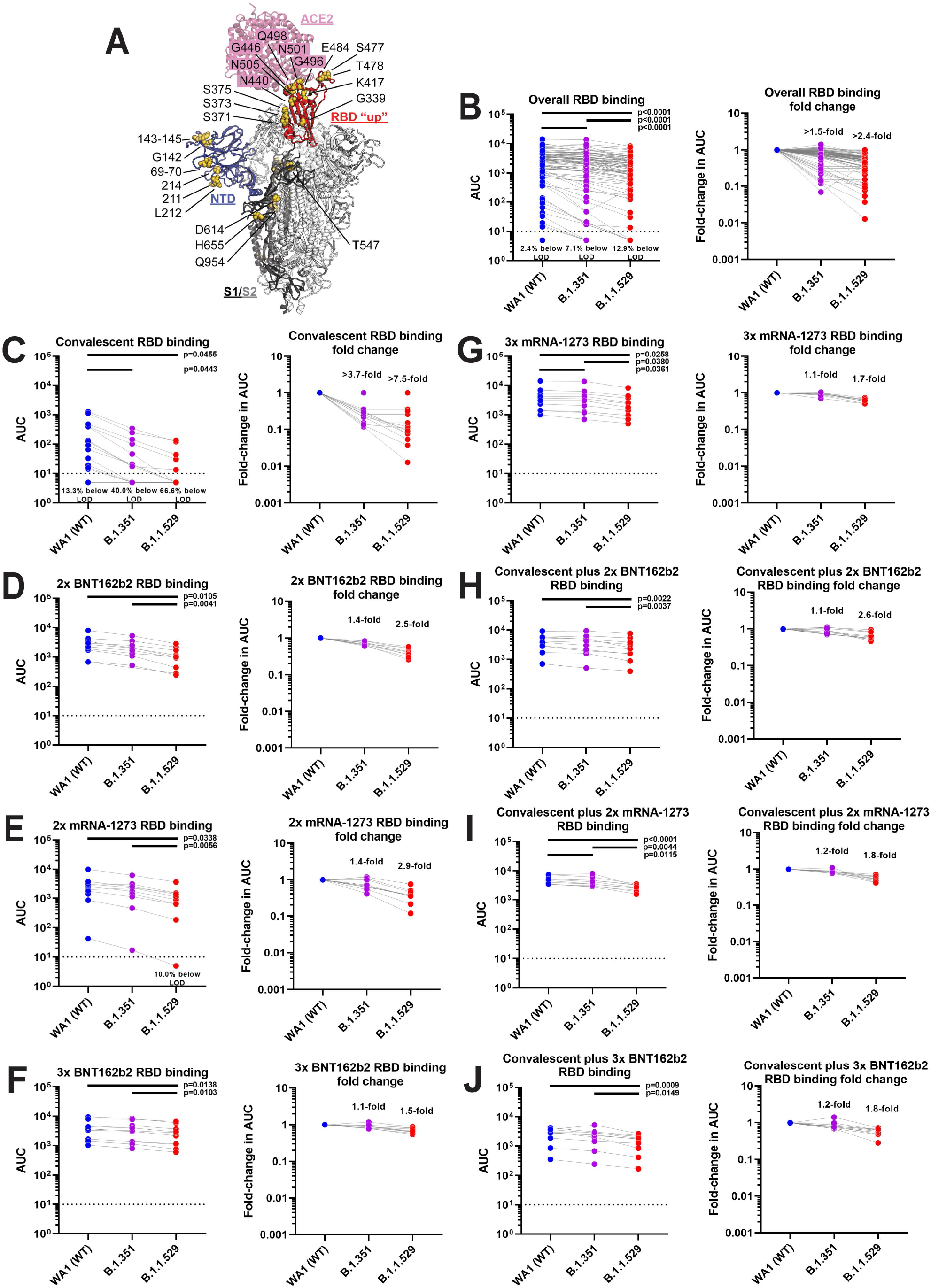
Sera of vaccinated individuals mostly maintain binding to the B.1.1.529 RBD. **A** shows a model of the B.1.1.529 spike protein in complex with the angiotensin converting enzyme 2 (ACE2) receptor with B.1.1.529 specific mutations indicated. The figure is bases on PDB 6M0J^24^ and 7C2L^25^ and was build in PyMol. **B** shows absolute titers (left) and fold reduction (right) for the combined samples, **C** to **J** shows the different groups. A one-way ANOVA with Tukey’s multiple comparisons test was used to compare the neutralization titers and significant p values (<0.05) are indicated in the figure. The exception is panel **D** where a mixed effects model had to be used due to a missing data point.

In addition to the RBD, the NTD is a prime target for B-cells after COVID-19 mRNA vaccination.^14^ The NTD also hosts neutralizing epitopes within and outside of the immunodominant ‘super site’. The NTD of B.1.1.529 carries a large number of amino acid substitutions, three deletions and one three amino acid long insertion which are, collectively, predicted to significantly change the ‘super site’ as well as neutralizing epitopes outside of the ‘super site’. To determine if infection induced and vaccine induced antibodies retain binding to the B.1.1.529 NTD, we expressed both the wild type and variant NTDs to probed by ELISA using the same 85 samples testing for neutralization. Surprisingly, binding to the NTD was maintained with relatively minor reductions (maximum 1.9-fold), suggesting either maintained binding (e.g. at lower affinity) to the ‘super site’ or the presence of a large number of unchanged epitopes within this domain (**Figure 3**).

**Figure 3:**
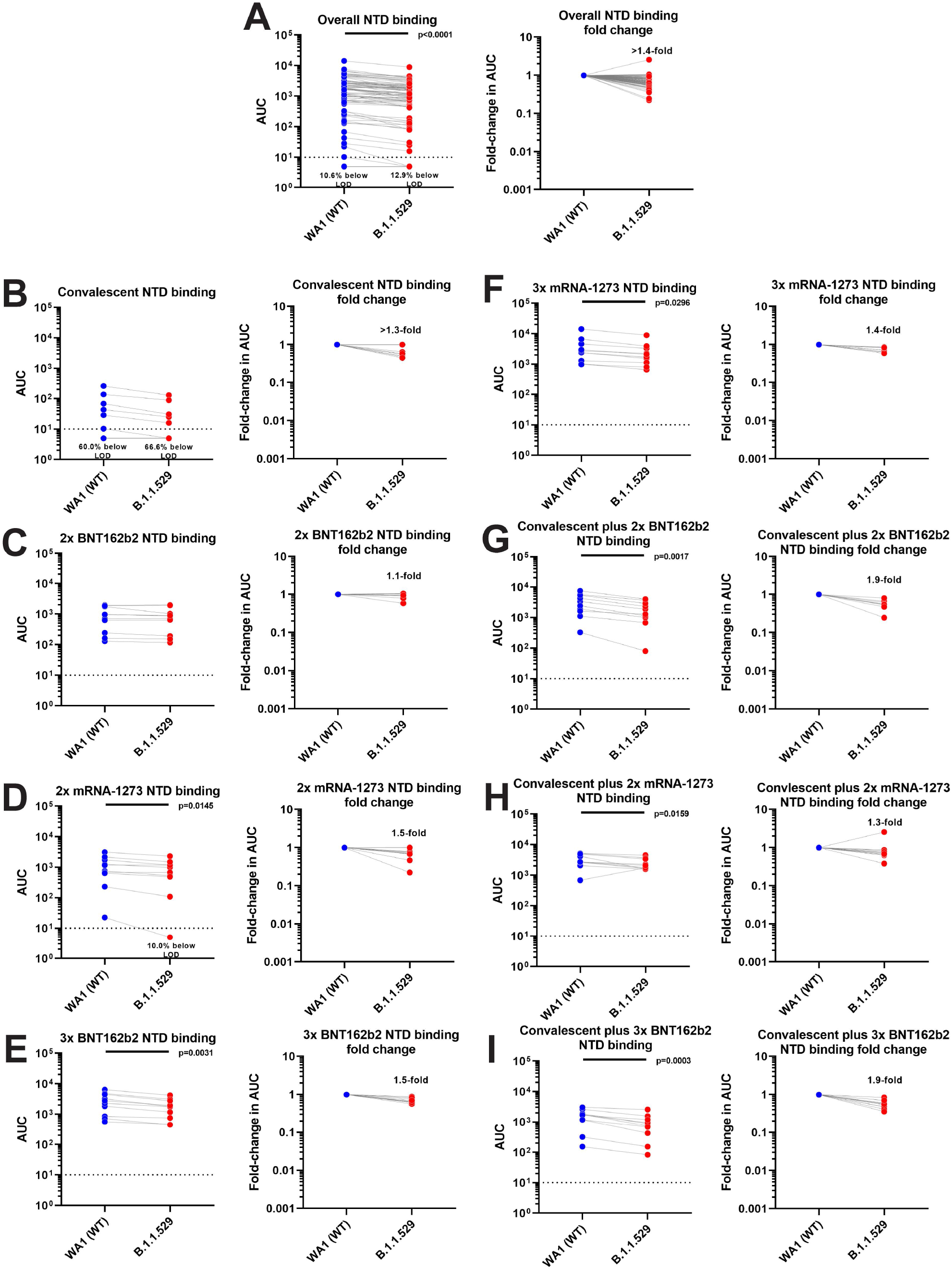
Serum of vaccinated individuals maintains binding to the B.1.1.529 NTD. **A** shows absolute titers (left) and fold reduction (right) for the combined samples, **B** to **I** shows the different groups. A student’s t test was used for comparing wild type and B.1.1.529 NTD binding data and significant p values (<0.05) are indicated in the figure.

Our data aligns well with initial reports on the impact of B.1.1.529 on *in vitro* neutralizing activity of convalescent and vaccine serum and expands on these initial reports by inclusion of subcohorts with divergent SARS-CoV-2 exposure history including infection induced, primary vaccine regimen as well as booster induced and hybrid immunity.^2,5,6^ We found that neutralizing activity of B.1.1.529 is most impacted in unvaccinated, convalescent individuals and in naïve individuals who acquired immunity through two mRNA COVID-19 vaccine doses. Our findings support recent reports describing significantly reduced protection from reinfection^15^ and basically non-existing vaccine effectiveness against symptomatic disease after two BNT162b2 vaccinations.^4^ However, boosted individuals had, at least, within the short time after the booster dose, significant protection against symptomatic disease in the range of 75%.^4^ While it is unclear how long this protection lasts, we observe titers similar to those in boosted individuals in convalescent vaccinated individuals, suggesting that those individuals may experience significant protection.

This study also provides first insights into B.1.1.529 RBD and NTD specific binding changes. Compared to the changes in neutralizing activities, binding was surprisingly well preserved especially against NTD, in general, and against the RBD in vaccinated, boosted and convalescent vaccinated individuals. It is conceivable, that these binding antibodies which often have non-neutralizing phenotypes in cell culture, contribute to protection from disease as has been seen for other viral infections. ^9-11^ In addition, the presence of strong binding antibodies suggest that, while some antibodies may have lost affinity for the drifted epitopes, B-cells may be recalled when encountering B.1.1.519 spike through infection or vaccination. This could lead to a strong anamnestic response, which could positively impact COVID-19 progression. It could also lead to the recruitment of these B-cells into germinal centers for further affinity maturation resulting in potent, high affinity neutralizing antibodies against B.1.1.529.^16^ Importantly, our data adds to the growing body of evidence suggesting that B.1.1.529 specific vaccines are urgently needed.

## Materials and methods

### Human Serum Samples

Convalescent and post-vaccine sera were collected from participants in the longitudinal observational PARIS (Protection Associated with Rapid Immunity to SARS-CoV-2) study.^8,17^ This cohort follows health care workers longitudinally since April 2020. The study was reviewed and approved by the Mount Sinai Hospital Institutional Review Board (IRB-20-03374). All participants signed written consent forms prior to sample and data collection. All participants provided permission for sample banking and sharing.

For the antigenic characterization of the antigenically diverse B.1.1.529 variant, we selected 85 serum samples from 54 participants. 20/54 participants were seronegative prior to vaccination while 34/54 had COVID-19 prior to vaccination (see **Supplemental Tables 1 and 2** for demographics and vaccine information). All participants with pre-vaccination immunity were infected in 2020 when only ancestral SARS-CoV-2 strains circulated in the New York metropolitan area. Convalescent samples (N=15) were obtained within three months of SARS-CoV-2 infection (average: 58 days, range: 23-87 days) while the post vaccinations samples were collected, on average, 23 days (range: 14-39 days) after the second dose (N= 40, 20 Pfizer 2x and 20 Moderna 2x) or 19 days (range: 14-33 days) after the third booster (N= 30, 20 Pfizer 3x and 10 Moderna 3x) vaccine dose.

### Cells

Vero-E6 cells expressing TMPRSS2 were cultured in Dulbecco’s modified Eagles medium (DMEM; Corning, #10-013-CV) containing 10% heat-inactivated fetal bovine serum (FBS; GeminiBio, #100-106) and 1% minimum essential medium (MEM) amino acids solution (Gibco, #11130051), supplemented with 100 U/ml penicillin and 100 μg/ml streptomycin (Gibco, #15140122), 100 μg/ml normocin (InvivoGen, #ant-nr), and 3 μg/ml puromycin (InvivoGen, #ant-pr). FreeStyle™ 293-F cells (Gibco, #R79007) were cultured in ESF-SFM medium (Expression Systems, cat. no. 98-001) supplemented with 100 U/ml penicillin and 100 μg/ml streptomycin (Gibco, #15140122). Expi293F™ Cells (Gibco, #A14527) were cultured in Expi293™ Expression Medium (Gibco, #A1435102) supplemented with 100 U/ml penicillin and 100 μg/ml streptomycin (Gibco, #15140122).

### Selection and culture of replication competent SARS-CoV-2 isolates

The Mount Sinai Pathogen Surveillance program (IRB approved, HS#13-00981) actively screens nasopharyngeal swab specimens from patients seeking care at the Mount Sinai Health System for emerging viral variants. After completion of the diagnostics, de-identified biospecimen were sequenced either using an established complete virus genome sequencing approach^18^ (e.g., Beta isolate USA/NY-MSHSPSP-PV27007/2021) or based on the spike S1 mutational profile determined by Spike-ID (Omicron, manuscript in preparation). The B.1.1.529 isolate USA/NY-MSHSPSP-PV44488/2021 represents one of the first cases diagnosed in New York State (female, age bracket: 30-40 years, mild COVID-19 symptoms, vaccinated and boosted) in late November 2021. The SARS-CoV-2 virus USA-WA1/2020 was used as wild-type reference (BEI resources, NR-52281). **Supplemental Table 3** summarizes the amino acid substitutions, insertions and deletions in the spike region of each of the three viral isolates

Viruses were grown by adding 200ul of viral transport media from the nasopharyngeal swabs to Vero-E6-TMPRSS2 cells in culture media supplemented with 0.5 μg/ml amphotericin B (Gibco, # 15290-018). Cytopathic effects (CPE) appears within 4-6 days at which point the culture supernatants was clarified by centrifugation at 4,000 g for 5 minutes. Expanded viral stocks used were sequence-verified and titered by the 50% tissue culture infectious dose (TCID_50_) method on Vero-E6-TMPRSS2 cells prior to use in micro neutralization assays.

### Generation of recombinant variant RBD and NTD proteins

The recombinant RBD proteins were produced using Expi293F cells (Life Technologies). The proteins were cloned into a mammalian expression vector, pCAGGS as described earlier^19,20^ and purified after transient transfections with each respective plasmid. Six-hundred million Expi293F cells were transfected using the ExpiFectamine 293 Transfection Kit and purified DNA. Supernatants were collected on day four post transfection, centrifuged at 4,000 g for 20 minutes and finally filtered using a 0.22 µm filter. Ni-nitrilotriacetic acid (Ni-NTA) agarose (Qiagen) was used to purify the proteins via gravity flow and proteins were eluted as previously described.^19,20^ The buffer was exchanged using Amicon centrifugal units (EMD Millipore) and all recombinant proteins were finally re-suspended in phosphate buffered saline (PBS). Proteins were also run on a sodium dodecyl sulphate (SDS) polyacrylamide gels (5–20% gradient; Bio-Rad) to check for purity.^21,22^ The NTD protein constructs (residues 1-306) were cloned into pVRC8400 expression vector between *Sal*I and *Not*I endonuclease restriction sites yielding an NTD with an human rhinovirus (HRV) 3C protease-cleavable C-terminal hexahistidine and a streptavidin-binding protein tags. The NTDs were transiently expressed in FreeStyle™ 293-F cells. Four days post-transfection, supernatants were harvested by centrifugation and further purified using immobilized metal affinity chromatography (IMAC) with cobalt-TALON® resin (Takara) followed by Superdex 200 Increase 10/300 GL size exclusion column (GE Healthcare).

### Enzyme linked immunosorbent assay (ELISA)

Antibody titers in sera from convalescent individuals and vaccinees were measured by a research grade ELISA using recombinant versions of the RBD and NTD of wild type SARS-CoV-2 as well as the B.1.351 (Beta), and B.1.1.529 (Omicron) (see **Supplemental Table 4** for specific substitutions in each variant). All samples were analyzed in a blinded manner. Briefly, 96-well microtiter plates (Corning, #353227) were coated with 50 μl/well of recombinant protein (2 μg/ml) overnight at 4 °C. Plates were washed three times with phosphate-buffered saline (PBS; Gibco, #10010-031) supplemented with 0.1% Tween-20 (PBS-T; Fisher Scientific ref. 202666) using an automatic plate washer (BioTek 405TS microplate washer). For blocking, PBS-T containing 3% milk powder (American Bio, # AB1010901000) was used. After 1-hour incubation at room temperature (RT), blocking solution was removed and initial dilutions (1:100) of heat-inactivated sera (in PBS-T 1%-milk powder) were added to the plates, followed by 2-fold serial dilutions. After 2-hour incubation, plates were washed three times with PBS-T and 50 μl/well of the pre-diluted secondary antibody anti-human IgG (Fab-specific) horseradish peroxidase (HRP) antibody (produced in goat; Sigma-Aldrich, Cat# A0293, RRID: AB_257875) diluted 1:3,000 in PBS-T containing 1% milk powder were added. After 1-hour incubation at RT, plates were washed three times with PBS-T and SigmaFast o-phenylenediamine dihydrochloride (Sigmafast OPD; Sigma-Aldrich, Ref. P9187-50SET) was added (100 μl/well) for 10min, followed by addition of 50 μl/well of 3 M hydrochloric acid (Thermo Fisher, Ref. S25856) to stop the reaction. Optical density was measured at a wavelength of 490 nm using a plate reader (BioTek, SYNERGY H1 microplate reader). The area under the curve (AUC) values were calculated and plotted using Prism 9 software (GraphPad).

### SARS-CoV-2 multi-cycle Microneutralization assay

Sera from vaccinees were used to assess the neutralization of wild type (WA1), B.1.351 (Beta) and B.1.1.529 (Omicron) SARS-CoV-2 isolates (**Supplementary Table 3**). All procedures were performed in a biosafety level 3 (BSL-3) facility at the Icahn School of Medicine at Mount Sinai following standard safety guidelines. Vero-E6-TMPRSS2 cells were seeded in 96-well high binding cell culture plates (Costar, #07620009) at a density of 20,000 cells/well in complete Dulbecco’s modified Eagle medium (cDMEM) one day prior to the infection. Heat inactivated serum samples (56°C for 1 hour) were serially diluted (3-fold) in minimum essential media (MEM; Gibco, #11430-030) supplemented with 2 mM L-glutamine (Gibco, #25030081), 0.1% sodium bicarbonate (w/v, HyClone, #SH30033.01), 10 mM 4-(2-hydroxyethyl)-1-piperazineethanesulfonic acid (HEPES; Gibco, #15630080), 100 U/ml penicillin, 100 μg/ml streptomycin (Gibco, #15140122) and 0.2% bovine serum albumin (BSA, MP Biomedicals, Cat#. 810063) starting at 1:10. Remdesivir (Medkoo Bioscience inc., #329511) was included to monitor assay variation. Serially diluted sera were incubated with 10,000 TCID_50_ of WT USA-WA1/2020 SARS-CoV-2, MSHSPSP-PV27007/2021 (B.1.351, Beta) or USA/NY-MSHSPSP-PV44488/2021 (B.1.1.529, Omicron) for one hour at RT, followed by the transfer of 120μl of the virus-sera mix to Vero-E6-TMPRSS2 plates. Infection proceeded for one hour at 37°C and inoculum was removed. 100 μl/well of the corresponding antibody dilutions plus 100μl/well of infection media supplemented with 2% fetal bovine serum (FBS; Gibco, #10082-147) were added to the cells. Plates were incubated for 48h at 37°C followed by fixation overnight at 4°C in 200 μl/well of a 10% formaldehyde solution. For staining of the nucleoprotein, formaldehyde solution was removed, and cells were washed with PBS (pH 7.4) (Gibco, #10010-031) and permeabilized by adding 150 μl/well of PBS, 0.1% Triton X-100 (Fisher Bioreagents, #BP151-100) for 15 min at RT. Permeabilization solution was removed, plates were washed with 200 μl/well of PBS (Gibco, #10010-031) twice and blocked with PBS, 3% BSA for 1 hour at RT. During this time the primary antibody was biotinylated according to manufacturer protocol (Thermo Scientific EZ-Link NHS-PEG4-Biotin). Blocking solution was removed and 100 μl/well of biotinylated mAb 1C7C7^23^, a mouse anti-SARS nucleoprotein monoclonal antibody generated at the Center for Therapeutic Antibody Development at The Icahn School of Medicine at Mount Sinai ISMMS (Millipore Sigma, Cat# ZMS1075) at a concentration of 1μg/ml in PBS, 1% BSA was added for 1 hour at RT. Cells were washed with 200 μl/well of PBS twice and 100 μl/well of HRP-conjugated streptavidin (Thermo Fisher Scientific) diluted in PBS, 1% BSA were added at a 1:2,000 dilution for 1 hour at RT. Cells were washed twice with PBS, and 100 μl/well of o-phenylenediamine dihydrochloride (Sigmafast OPD; Sigma-Aldrich) were added for 10 min at RT, followed by addition of 50 μl/well of a 3 M HCl solution (Thermo Fisher Scientific). Optical density (OD) was measured (490 nm) using a microplate reader (Synergy H1; Biotek). Analysis was performed using Prism 7 software (GraphPad). After subtraction of background and calculation of the percentage of neutralization with respect to the “virus only” control, a nonlinear regression curve fit analysis was performed to calculate the 50% inhibitory dilution (ID_50_), with top and bottom constraints set to 100% and 0% respectively. All samples were analyzed in a blinded manner.

### Statistics

A one-way ANOVA with Tukey’s multiple comparisons test was used to compare the neutralization and RBD binding antibody titers. The exception is the 2x BNT162b2 RBD ELISA group where a mixed effects model had to be used due to a missing data point. A student’s t test was used for comparing wild type and B.1.1.529 NTD binding data. Statistical analyses were performed using Prism 9 software (GraphPad).

## Data Availability

All data produced in the present study are available upon reasonable request to the authors

## Acknowledgements

We thank the study participants for their generosity and willingness to participate in longitudinal COVID-19 research studies. None of this work would be possible without their contributions.

We would like to thank Dr. Randy A. Albrecht for oversight of the conventional BSL3 biocontainment facility, which makes our work with live SARS-CoV-2 possible. Furthermore, we would like to thank Drs. Janine Kimpel and Dorothee von Laer for their willingness to speedily share B.1.1.529 virus isolates (even though they could not included in this work). We are also grateful for Mount Sinai’s leadership during the COVID-19 pandemic. We want to especially thank Drs. Peter Palese, Carlos Cordon-Cardo, Dennis Charney, David Reich and Kenneth Davis for their support and Daniel Caughey for expert administrative assistance.

This work is part of the PARIS/SPARTA studies funded by the NIAID Collaborative Influenza Vaccine Innovation Centers (CIVIC) contract 75N93019C00051. In addition, this work was also partially funded by the NIAID Centers of Excellence for Influenza Research and Response (CEIRR) contract and 75N93021C00014 by anonymous donors. This work is part of the NIAID SARS-CoV-2 Assessment of Viral Evolution (SAVE) program.

## Supplemental Tables

**Supplemental Table 1:** Overall description of samples used.

|  | COVID-19 | Two vaccine doses |  |  |  | Three vaccine doses |  |  |
| --- | --- | --- | --- | --- | --- | --- | --- | --- |
|  | Convalescent | BNT16 2b2 |  | mRNA-1273 |  | BNT16 2b2 |  | mRNA-1273 |
|  |  | naïve | convalescent | naïve | convalescent | naïve | convalescent | naïve |
| **Total N** | 15 | 10 | 10 | 10 | 10 | 10 | 10 | 10 |
| **Sex** |  |  |  |  |  |  |  |  |
| Female | 10 | 8 | 7 | 8 | 5 | 8 | 7 | 8 |
| Male | 5 | 2 | 3 | 2 | 5 | 2 | 3 | 2 |
| **Days since infection** |  |  |  |  |  |  |  |  |
| Average (days) | 58 | no infection |  | no infection |  | no infection |  | no infection |
| Range (days) | 23-87 | no infection |  | no infection |  | no infection |  | no infection |
| **Days since vaccine dose** |  |  |  |  |  |  |  |  |
| Average (days) | no vaccine | 18 | 26 | 26 | 20 | 19 | 20 | 19 |
| Range (days) | no vaccine | 14-21 | 15-39 | 14-36 | 15-28 | 14-28 | 14-30 | 14-33 |

**Supplemental Table 2:** Detailed description of samples used. Summary of the metadata of the 54 PARIS participants from whom a total of 85 serum samples were analyzed. For 31 PARIS participants, samples from two different time points were included (Post-Vax, Post-Boost or Post-Infection, Post-Vax). The following abbreviations are used in the table: Post-infection: serum collected 23 to 87 days after SARS-CoV-2 diagnosis Post-Vax: serum collected 14-39 days after the second dose of mRNA vaccine Post-Boost: serum collected 14-30 days after the booster vaccine dose no vax: participant was not vaccinated at the time of sample collection n.a: data not available

| Participant ID | Age Bracket | Sex | Ancestry | Time points included in this study | SARS-CoV-2 infection prior to vaccination | Vaccine Type |
| --- | --- | --- | --- | --- | --- | --- |
| OMI-001 | 30-39 | Female | Caucasian | Post-Vax, Post-Boost | No | Moderna |
| OMI-002 | 18-29 | Female | Caucasian | Post-Vax, Post-Boost | No | Moderna |
| OMI-003 | 40-49 | Male | Caucasian | Post-Vax, Post-Boost | No | Moderna |
| OMI-004 | 40-49 | Female | Caucasian | Post-Vax, Post-Boost | No | Moderna |
| OMI-005 | 40-49 | Female | Caucasian | Post-Vax, Post-Boost | No | Moderna |
| OMI-006 | 40-49 | Male | Caucasian | Post-Vax, Post-Boost | No | Moderna |
| OMI-007 | 50-59 | Female | Caucasian | Post-Vax, Post-Boost | No | Moderna |
| OMI-008 | 60-69 | Female | Asian | Post-Vax, Post-Boost | No | Moderna |
| OMI-009 | 30-39 | Female | Caucasian | Post-Vax, Post-Boost | No | Moderna |
| OMI-010 | 18-29 | Female | Caucasian | Post-Vax, Post-Boost | No | Moderna |
| OMI-011 | 40-49 | Female | Other | Post-Vax, Post-Boost | No | Pfizer |
| OMI-012 | 18-29 | Female | Asian | Post-Vax, Post-Boost | No | Pfizer |
| OMI-013 | 60-69 | Female | Caucasian | Post-Vax, Post-Boost | No | Pfizer |
| OMI-014 | 50-59 | Female | Caucasian | Post-Vax, Post-Boost | No | Pfizer |
| OMI-015 | 18-29 | Female | Caucasian | Post-Vax, Post-Boost | No | Pfizer |
| OMI-016 | 70-79 | Male | Caucasian | Post-Vax, Post-Boost | No | Pfizer |
| OMI-017 | 30-39 | Female | Asian Indian | Post-Vax, Post-Boost | No | Pfizer |
| OMI-018 | 30-39 | Female | Caucasian | Post-Vax, Post-Boost | No | Pfizer |
| OMI-019 | 40-49 | Male | Caucasian | Post-Vax, Post-Boost | No | Pfizer |
| OMI-020 | 40-49 | Female | n.a. | Post-Vax, Post-Boost | No | Pfizer |
| OMI-021 | 40-49 | Female | Asian | Post-Infection, Post-Vax | Yes | Pfizer |
| OMI-022 | 30-39 | Female | Caucasian | Post-Infection, Post-Vax | Yes | Pfizer |
| OMI-023 | 30-39 | Female | Asian | Post-Infection, Post-Vax | Yes | Pfizer |
| OMI-024 | 30-39 | Male | Caucasian | Post-Infection, Post-Vax | Yes | Pfizer |
| OMI-028 | 18-29 | Female | Caucasian | Post-Infection, Post-Vax | Yes | Moderna |
| OMI-029 | 30-39 | Female | Caucasian | Post-Infection, Post-Vax | Yes | Pfizer |
| OMI-032 | 50-59 | Male | Caucasian | Post-Infection, Post-Vax | Yes | Pfizer |
| OMI-027 | 40-49 | Female | Caucasian | Post-Infection, Post-Vax | Yes | Pfizer |
| OMI-033 | 30-39 | Female | Caucasian | Post-Infection, Post-Vax | Yes | Pfizer |
| OMI-034 | 40-49 | Male | Latino | Post-Infection, Post-Vax | Yes | Pfizer |
| OMI-035 | 30-39 | Female | Caucasian | Post-Infection, Post-Vax | Yes | Pfizer |
| OMI-025 | 50-59 | Male | Latino | Post-Infection | Yes | no vax |
| OMI-026 | 30-39 | Male | Caucasian | Post-Infection | Yes | no vax |
| OMI-030 | 40-49 | Female | Other | Post-Infection | Yes | no vax |
| OMI-031 | 30-39 | Female | Mexican | Post-Infection | Yes | no vax |
| OMI-036 | 50-59 | Male | Asian | Post-Vax | Yes | Moderna |
| OMI-037 | 30-39 | Male | Caucasian | Post-Vax | Yes | Moderna |
| OMI-038 | 18-29 | Male | Caucasian | Post-Vax | Yes | Moderna |
| OMI-039 | 30-39 | Female | Caucasian | Post-Vax | Yes | Moderna |
| OMI-040 | 60-69 | Female | African American | Post-Vax | Yes | Moderna |
| OMI-041 | 40-49 | Female | Caucasian | Post-Vax | Yes | Moderna |
| OMI-042 | 40-49 | Female | Caucasian | Post-Vax | Yes | Moderna |
| OMI-043 | 60-69 | Male | Caucasian | Post-Vax | Yes | Moderna |
| OMI-044 | 40-49 | Male | n.a. | Post-Vax | Yes | Moderna |
| OMI-045 | 30-39 | Male | Caucasian | Post-Boost | Yes | Pfizer |
| OMI-046 | 30-39 | Female | Caucasian | Post-Boost | Yes | Pfizer |
| OMI-047 | 50-59 | Female | n.a. | Post-Boost | Yes | Pfizer |
| OMI-048 | 60-69 | Male | Caucasian | Post-Boost | Yes | Pfizer |
| OMI-049 | 60-69 | Male | Caucasian | Post-Boost | Yes | Pfizer |
| OMI-050 | 30-39 | Female | African American | Post-Boost | Yes | Pfizer |
| OMI-051 | 50-59 | Female | Caucasian | Post-Boost | Yes | Pfizer |
| OMI-052 | 50-59 | Female | Caucasian | Post-Boost | Yes | Pfizer |
| OMI-053 | 18-29 | Female | Caucasian | Post-Boost | Yes | Pfizer |
| OMI-054 | 30-39 | Female | Caucasian | Post-Boost | Yes | Pfizer |

**Supplementary Table 3:**
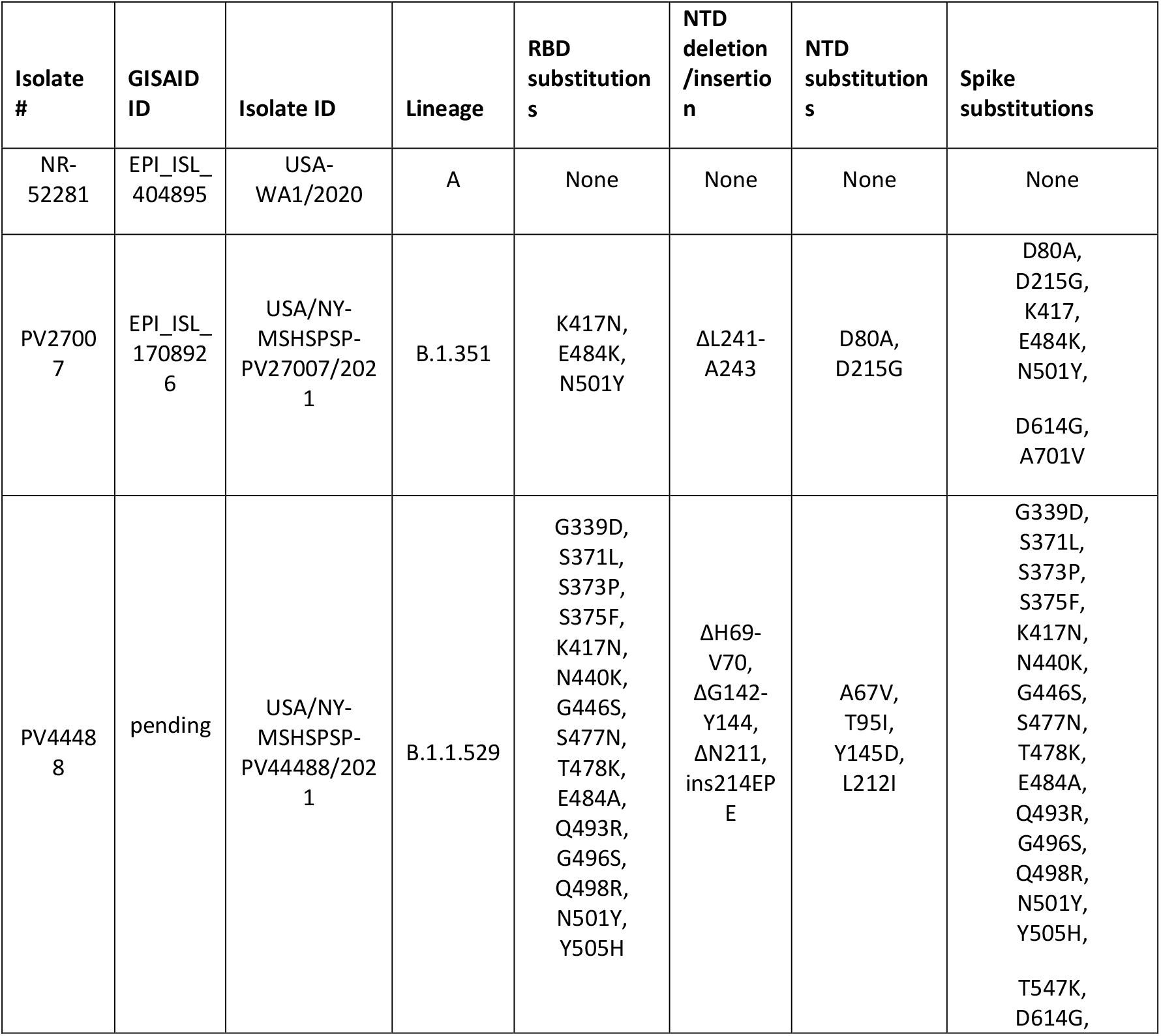
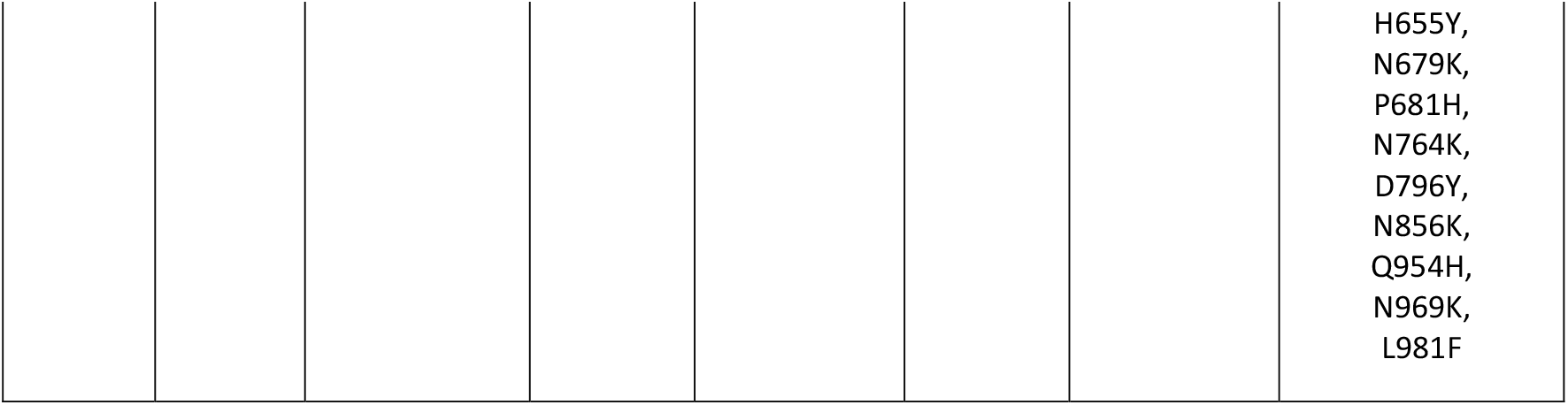
Information on the viral isolates used in neutralization assays.

**Supplemental Table 4:** Overview of the mutations encoded in the RBD and NTD proteins used for the binding assays.

| Mutations in the sequence of recombinant RBD and spike proteins generated for this study |  |  |  |  |  |
| --- | --- | --- | --- | --- | --- |
| Protein | Lineage | RBD substitutions | NTD deletions | NTD insertions | NTD substitutions |
| RBD | WA1 | None | NA | NA | NA |
| RBD | B.1.351 | K417K, E484K, N501Y | NA | NA | NA |
| RBD | B.1.1.529 | G339D, S371L, S373P,<br>S375F, K417N, N440K,<br>G446S, S477N, T478K,<br>E484A, G496S, Q498R,<br>N501Y, Y505H | NA | NA | NA |
| NTD | WA1 | NA | None | None | None |
| NTD | B.1.1.529 | NA | 69-70, 143-145,<br>211 | 214EPE | A67V, T95I, G142D, L212I |

## References

1 Gu, H. et al. Probable Transmission of SARS-CoV-2 Omicron Variant in Quarantine Hotel, Hong Kong, China, November 2021. Emerg Infect Dis 28, doi:10.3201/eid2802.212422 (2021).

2 Cele, S. et al. SARS-CoV-2 Omicron has extensive but incomplete escape of Pfizer BNT162b2 elicited neutralization and requires ACE2 for infection. medRxiv, 2021.2012.2008.21267417, doi:10.1101/2021.12.08.21267417 (2021).

3 Brandal, L. T. et al. Outbreak caused by the SARS-CoV-2 Omicron variant in Norway, November to December 2021. Eurosurveillance 26, 2101147, doi:doi:https://doi.org/10.2807/1560-7917.ES.2021.26.50.2101147 (2021).

4 Andrews, N. et al. Effectiveness of COVID-19 vaccines against the Omicron (B.1.1.529) variant of concern. medRxiv, 2021.2012.2014.21267615, doi:10.1101/2021.12.14.21267615 (2021).

5 Wilhelm, A. et al. Reduced Neutralization of SARS-CoV-2 Omicron Variant by Vaccine Sera and Monoclonal Antibodies. medRxiv, 2021.2012.2007.21267432, doi:10.1101/2021.12.07.21267432 (2021).

6 Rössler, A., Riepler, L., Bante, D., Laer, D.v. & Kimpel, J. SARS-CoV-2 B.1.1.529 variant (Omicron) evades neutralization by sera from vaccinated and convalescent individuals. medRxiv, 2021.2012.2008.21267491, doi:10.1101/2021.12.08.21267491 (2021).

7 Uriu, K. et al. Neutralization of the SARS-CoV-2 Mu Variant by Convalescent and Vaccine Serum. N Engl J Med 385, 2397–2399, doi:10.1056/NEJMc2114706 (2021).

8 Carreño, J. M. et al. Evidence for retained spike-binding and neutralizing activity against emerging SARS-CoV-2 variants in serum of COVID-19 mRNA vaccine recipients. EBioMedicine 73, 103626, doi:10.1016/j.ebiom.2021.103626 (2021).

9 Dilillo, D. J., Tan, G. S., Palese, P. & Ravetch, J. V. Broadly neutralizing hemagglutinin stalk-specific antibodies require FcγR interactions for protection against influenza virus in vivo. Nat Med, doi:10.1038/nm.3443 (2014).

10 Asthagiri Arunkumar, G. et al. Broadly Cross-Reactive, Nonneutralizing Antibodies against Influenza B Virus Hemagglutinin Demonstrate Effector Function-Dependent Protection against Lethal Viral Challenge in Mice. J Virol 93, doi:10.1128/JVI.01696-18 (2019).

11 Ng, S. et al. Novel correlates of protection against pandemic H1N1 influenza A virus infection. Nat Med 25, 962–967, doi:10.1038/s41591-019-0463-x (2019).

12 Gilbert, P. B. et al. Immune correlates analysis of the mRNA-1273 COVID-19 vaccine efficacy clinical trial. Science, eab3435 (2021).

13 Earle, K. A. et al. Evidence for antibody as a protective correlate for COVID-19 vaccines. Vaccine, doi:10.1016/j.vaccine.2021.05.063 (2021).

14 Amanat, F. et al. SARS-CoV-2 mRNA vaccination induces functionally diverse antibodies to NTD, RBD, and S2. Cell, doi:10.1016/j.cell.2021.06.005 (2021).

15 Pulliam, J. R. C. et al. Increased risk of SARS-CoV-2 reinfection associated with emergence of the Omicron variant in South Africa. medRxiv, 2021.2011.2011.21266068, doi:10.1101/2021.11.11.21266068 (2021).

16 Turner, J. S. et al. SARS-CoV-2 mRNA vaccines induce persistent human germinal centre responses. Nature, doi:10.1038/s41586-021-03738-2 (2021).

17 Krammer, F. et al. Antibody Responses in Seropositive Persons after a Single Dose of SARS-CoV-2 mRNA Vaccine. N Engl J Med, doi:10.1056/NEJMc2101667 (2021).

18 Gonzalez-Reiche, A. S. et al. Introductions and early spread of SARS-CoV-2 in the New York City area. Science, doi:10.1126/science.abc1917 (2020).

19 Amanat, F. et al. A serological assay to detect SARS-CoV-2 seroconversion in humans. Nat Med 26, 1033–1036, doi:10.1038/s41591-020-0913-5 (2020).

20 Stadlbauer, D. et al. SARS-CoV-2 Seroconversion in Humans: A Detailed Protocol for a Serological Assay, Antigen Production, and Test Setup. Curr Protoc Microbiol 57, e100, doi:10.1002/cpmc.100 (2020).

21 Margine, I., Palese, P. & Krammer, F. Expression of functional recombinant hemagglutinin and neuraminidase proteins from the novel H7N9 influenza virus using the baculovirus expression system. J Vis Exp, e51112, doi:10.3791/51112 (2013).

22 Amanat, F. et al. Antibodies to the Glycoprotein GP2 Subunit Cross-React between Old and New World Arenaviruses. mSphere 3, doi:10.1128/mSphere.00189-18 (2018).

23 Amanat, F. et al. An In Vitro Microneutralization Assay for SARS-CoV-2 Serology and Drug Screening. Curr Protoc Microbiol 58, e108, doi:10.1002/cpmc.108 (2020).

24 Lan, J. et al. Structure of the SARS-CoV-2 spike receptor-binding domain bound to the ACE2 receptor. Nature 581, 215–220, doi:10.1038/s41586-020-2180-5 (2020).

25 Chi, X. et al. A neutralizing human antibody binds to the N-terminal domain of the Spike protein of SARS-CoV-2. Science 369, 650–655, doi:10.1126/science.abc6952 (2020).

